# Anatomical dynamics define cancer cachexia subtypes and identify systemic inflammation as a marker of lethal wasting

**DOI:** 10.64898/2026.05.04.26352250

**Authors:** Sonia Boscenco, Venise Jan Castillon, Jamie Wang, Ethan Tse, Samuel S. Freeman, Ziad Bakouny, Saksham Mohan, X. Alex Guo, River Walser, Junmin Song, Constantinos P. Zambirinis, Linda Bojmar, Ritesh R. Kotecha, Marc Hilmi, Michael S. May, Gerardo A. Vitiello, Tobias Janowitz, Marcus D. Goncalves, Natalie Gangai, David Lyden, Adrianna Z. Herskovits, Puneeth Iyengar, William R. Jarnigan, Robert E. Schwartz, Ramon Sosa, Justin Jee, Eileen M. O’Reilly, Nikolaus Schultz, Sohrab P. Shah, Wungki Park, John W. Garrett, Perry J. Pickhardt, Nathaniel C. Swinburne, Ed Reznik

## Abstract

Cancer cachexia is a wasting syndrome that remodels the anatomy of the patient. How this remodeling unfolds across tissues, whether it defines distinct disease states, and how these states relate to underlying biology remain unknown. We used longitudinal computed tomography imaging from 4,516 patients to quantify evolution of muscle, adipose, and organs during cachexia. Across two independent institutional cohorts, unsupervised analysis identified three reproducible anatomical subtypes of cachexia, including an inflammatory Type A marked by progressive hepatosplenic enlargement and inferior survival, a Type B dominated by visceral organ atrophy, and a mild Type C. These anatomical subtypes were associated with distinct serological signatures and reflected in molecular phenotypes in tumors and non-cancerous liver tissue, establishing cachexia as discrete anatomical disease states that link whole-body remodeling to systemic and tissue-level biology. This anatomy-first framework for cachexia classification provides a foundation for future patient stratification and development of subtype-specific anti-cachexia therapies.

## Introduction

The endurance and treatment of cancer affects the human body beyond the physical extent of the tumor. While the primary goal of cancer therapy is to treat the tumor to extend life, the progressive deterioration of the patient, arising equally from the systemic effects of tumor burden as well as the toxic effects of therapy, reduces the capacity to receive potentially life-saving therapy. And yet, while the morphologic and molecular phenotypes of tumors are deeply integrated into clinical care and cancer science, there is comparatively little measurement of non-involved, non-malignant tissue from cancer patients, leaving a reservoir of biological discovery potentially untapped.

The aim of this work is to study the evolution of the human body during periods of cancer cachexia (“cachexia”), an involuntary wasting syndrome associated with profound reductions in quality-of-life and overall survival that impacts 50-80% of all cancer patients^1–3^. Cachexia affects non-malignant muscle and adipose tissue which (unlike tumor tissue) is not subject to routine biopsy during clinical care. Consequently, a historically significant obstacle to studying cachexia in patients with cancer, rather than model systems, has been the difficulty of sampling and making measurements on non-malignant tissue, including but not limited to adipose and muscle. Recent advances in the automated analysis of routinely collected imaging data, especially abdominal computed tomography (CT) scans, have enabled the analysis of anatomical changes in muscle, adipose, and organs in cancer patients at-scale^4–9^. Here, we apply these computational methods to routinely-collected longitudinal CT scans on patients with cancer, enabling rich, scaled, and dynamic anatomical phenotyping of cancer patients across periods of clinically significant weight loss.

The conceptual framework of molecular subtypes of cancer, each of which exhibits a characteristic pattern of genetic, transcriptional, or immunologic features, has underpinned transformative biological discoveries and enabled the design of rationally-targeted therapeutics. Recently, molecular profiling of skeletal muscle from a small number of cachectic cancer patients provocatively suggested that cachexia emerges via one of potentially many subtypes^10^. Here, we discover clinically significant subtypes of cachexia derived from dynamic measurements of patient anatomy, rather than static measurements of molecular factors. We go on to demonstrate that anatomical subtypes capture the net effect of tumor burden, host metabolism, and systemic inflammation—features that are not confined to a single tissue and are likely to holistically shape clinical trajectories.

## Results

### Anatomical evolution across periods of weight loss

We sought to understand how the anatomy of cancer patients changes as they endure cachexia. To do so, we retrospectively identified 4,516 patients across 13 cancer types satisfying two criteria: (1) having experienced weight loss meeting the consensus weight loss criteria for cachexia that have been implemented in recent clinical trials of targeted agents against cachexia^11^ and (2) having received an abdominal contrast-enhanced CT scan within 90 days of initiation of weight loss, and within 90 days of cessation of weight loss (see **Methods**; **Figure 1a-b**). For each scan, we used a previously developed and automated AI pipeline (Opportunistic Screening Consortium in Abdominal Radiology, “OSCAR”, see **Methods**)^12–14)^ to quantify the area and radiographic density of subcutaneous adipose tissue (SAT), visceral adipose tissue (VAT), intramuscular adipose tissue (IMAT), and skeletal muscle (SKM) at the third-lumbar spinal region (L3), as well as the volume and radiographic density of the liver, spleen, kidney, and pancreas, and bone mineral density (BMD) (**Supplementary Table 1**). Thus, for each patient, at 2 distinct time points corresponding to pre- and post-cachexia, we obtained granular information on body composition. OSCAR has previously been validated and benchmarked on >500,000 geographically-diverse patients from the general population at multiple health systems^12,13,15,16^. To validate the accuracy of OSCAR on a population of individuals with cancer, we manually segmented the SKM, SAT, and VAT areas of CT scans from 100 colorectal adenocarcinoma patients at the L3 region. We observed a high degree of concordance between manual and automated segmentation for SAT (Pearson P < 2.2×10^-16^, r = 0.94), VAT (Pearson P < 2.2×10^-16^, r = 0.9), and SKM (Pearson P < 2.2×10^-16^, r = 0.96), confirming the accuracy of OSCAR in our cohort (**Figure 1c**).

**Figure 1:**
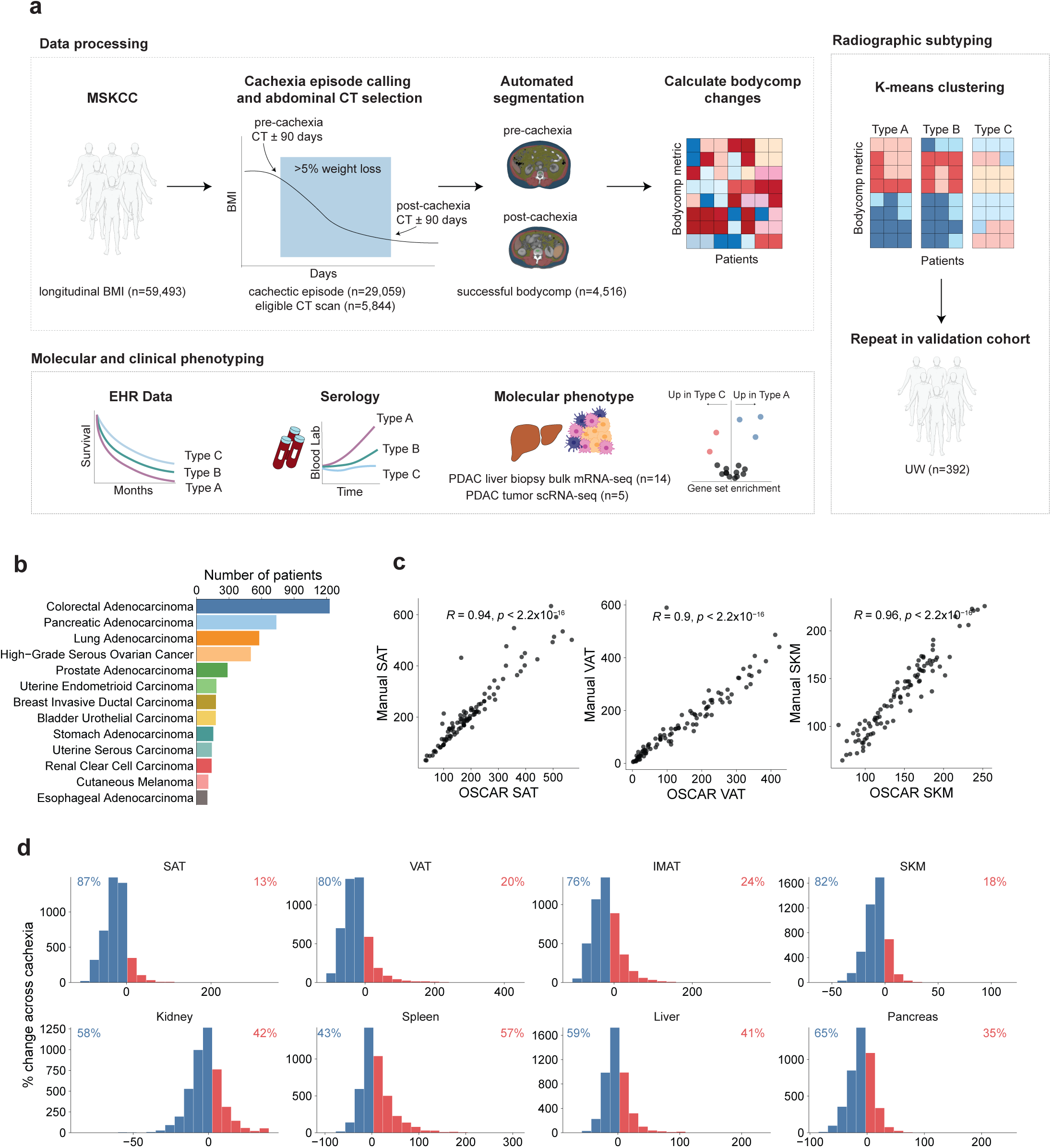
Landscape of body composition changes during cachexia. **a.** Design of study. Patients with weight loss meeting consensus weight loss criteria for cachexia and eligible scans flanking the weight loss period are analyzed for changes in body composition. Cachexia subtypes are identified and analyzed through clinical outcomes, serology, and molecular phenotypes. **b.** Cancer type distribution of patient cohort. **c.** Automatic OSCAR segmentations versus manual segmentations for a subset of 100 CRC patients for SAT (R = 0.94, p < 2.2×10-16), VAT (R = 0.90, p < 2.2×10-16), and SKM (R = 0.96, p < 2.2×10-16), respectively. R values and p-values derived from Pearson correlations. **d.** Histogram representation of changes across weight loss windows, represented as % changes for areas (cm2) and volumes (cm3). Annotated value represents percentage of patients with loss in that tissue or organ (blue), or gain (red).

To determine the physiological trajectory of cachectic wasting across the entire patient cohort, we calculated the percent change (for areas and volumes) and raw change (for densities) between the pre and post-cachexia scans at the axial slice corresponding to the L3 lumbar vertebra (**Figure 1d; Supplementary Figure 2a, Supplementary Table 2**). We repeated these quantifications using segmentations of spinal regions L1, T10, and T12 and determined that they were highly correlated with corresponding metrics at L3, consistent with prior literature describing body composition assessment at L3 as an accurate surrogate for systemic body composition changes (**Supplementary Figure 1a-c**)^6,17^.

We proceeded to examine how the bodies of patients with cancer evolve over time during cachexia. Changes in body composition were broadly distributed across all tissues and other organs. As expected, SAT, VAT, and SKM loss were the most prevalent changes, affecting 87%, 80%, and 82% of patients, respectively (**Fig 1d, Supplementary Figure 2a**). In contrast to adipose and muscle, patients demonstrated heterogeneous patterns of changes to organ volumes. For example, 65% of patients demonstrated volumetric loss of the pancreas, whereas progressive increases in spleen volume were evident in 57% of patients. To assess the robustness of cachectic anatomical changes, we analyzed cachectic episodes from a second, independent cohort of 392 patients with cancer from the University of Wisconsin-Madison (UW) with eligible CT scans flanking their weight-loss episodes. In the validation cohort, we confirmed that loss of SAT (79%), VAT (83%), and SKM (77%) were the dominant phenotypes of cachexia. We also recovered trends in key organ volumes, where pancreatic volume loss was highly prevalent (73%) and splenic volume enlargement affected a small majority of patients (51%) (**Supplementary Figure 4a**)

To assess the extent to which aspects of anatomical evolution are specific to cachectic episodes, rather than normal fluctuations during cancer treatment, we assembled an additional cohort of 280 patients with lung adenocarcinoma, uterine endometrioid carcinoma, pancreatic adenocarcinoma, and renal clear cell carcinoma who did not develop >5% weight loss at any time of their measurable disease course. Across 1,972 longitudinal time points, ranging from three to fifteen per patient spaced three to nine months apart, we quantified time-normalized changes in body composition to form a representation of normal variation of SAT, VAT, SKM, and organ volumes (see **Methods**). Compared to these non-cachectic controls, patients undergoing active weight loss exhibited significantly greater depletion of SAT (Welch two sample t-test p < 2.2×10^-16^, Cohen’s d = -0.881), VAT (Welch two sample t-test p < 2.2×10^-16^, Cohen’s d = -0.551), and SKM (Welch two sample t-test p < 2.2×10^-16^, Cohen’s d = -0.855) per month (**Supplementary Figure 2b**). Pancreas volume was significantly decreased in cachectic patients compared to non-cachectic controls (Welch two sample t-test p < 2.2×10^-16^, Cohen’s d = -0.345), whereas splenic volume was elevated in cachexia (Welch two sample t-test p < 2.2×10^-16^, Cohen’s d = 0.293). Significant changes were not detectable in either the liver (Welch two sample t-test p = 0.432, Cohen’s d = -0.0200) or the kidney (Welch two sample t-test p = 0.787, Cohen’s d = -0.00635) (**Supplementary Figure 2c**). These data confirm that radiographically derived body composition changes during cachexia, and in particular adipose and muscle, reflect physiological changes that are distinct from those in patients not demonstrating cachectic weight loss.

Historically, cachexia has been described to occur more commonly in a set of specific cancer types such as lung and pancreatic adenocarcinomas, although emerging systematic analyses of large cohorts of patients show that cachexia is common across many cancers^18^. We therefore tested whether anatomical evolution differed across cancer types. We fit a multivariate linear regression model and associated each body composition metric with cancer type, controlling for age, sex, and pre-cachectic BMI. This analysis identified 62 statistically significant associations between cancer type and a change in a single body composition metric (**Figure 2a, c-d, Supplementary Table 3**). SKM loss was not characteristically different in any given cancer type, whereas SAT loss was particularly pronounced in pancreatic adenocarcinoma (q = 3.74×10^-12^, β = -8.74), stomach adenocarcinoma (q = 2.73×10^-5^, β = -11.1), and esophageal adenocarcinoma (q = 2.46×10^-2^, β = -7.99), which are among the cancer subtypes canonically associated with cachexia development^19,20^. Additionally, we observed that several organ level changes were stronger in specific cancer types, such as spleen volume gain in colorectal adenocarcinoma (q = 1.28×10^-6^, β = 6.73) and pancreatic adenocarcinoma (q = 1.46×10^-7^, β = 8.86). To confirm the robustness of histology-specific anatomical changes, we repeated this analysis in the UW validation cohort. Histological subtype identity was derived from ICD codes, which enabled classification to the granularity of primary site location. We restricted our analysis to the disease with the largest sample size (pancreatic cancer, n=41), where we recapitulated key patterns, such as characteristically strong depletion of SAT area (q = 1.97×10^-4^, β = -17.9) and increase of SAT density (q = 1.44×10^-5^, β = 6.25) (**Figure 2b,e,f**). These data confirm that aspects of anatomical evolution are dictated by disease-specific factors but also identify patterns of anatomical evolution that transcend individual diseases.

**Figure 2:**
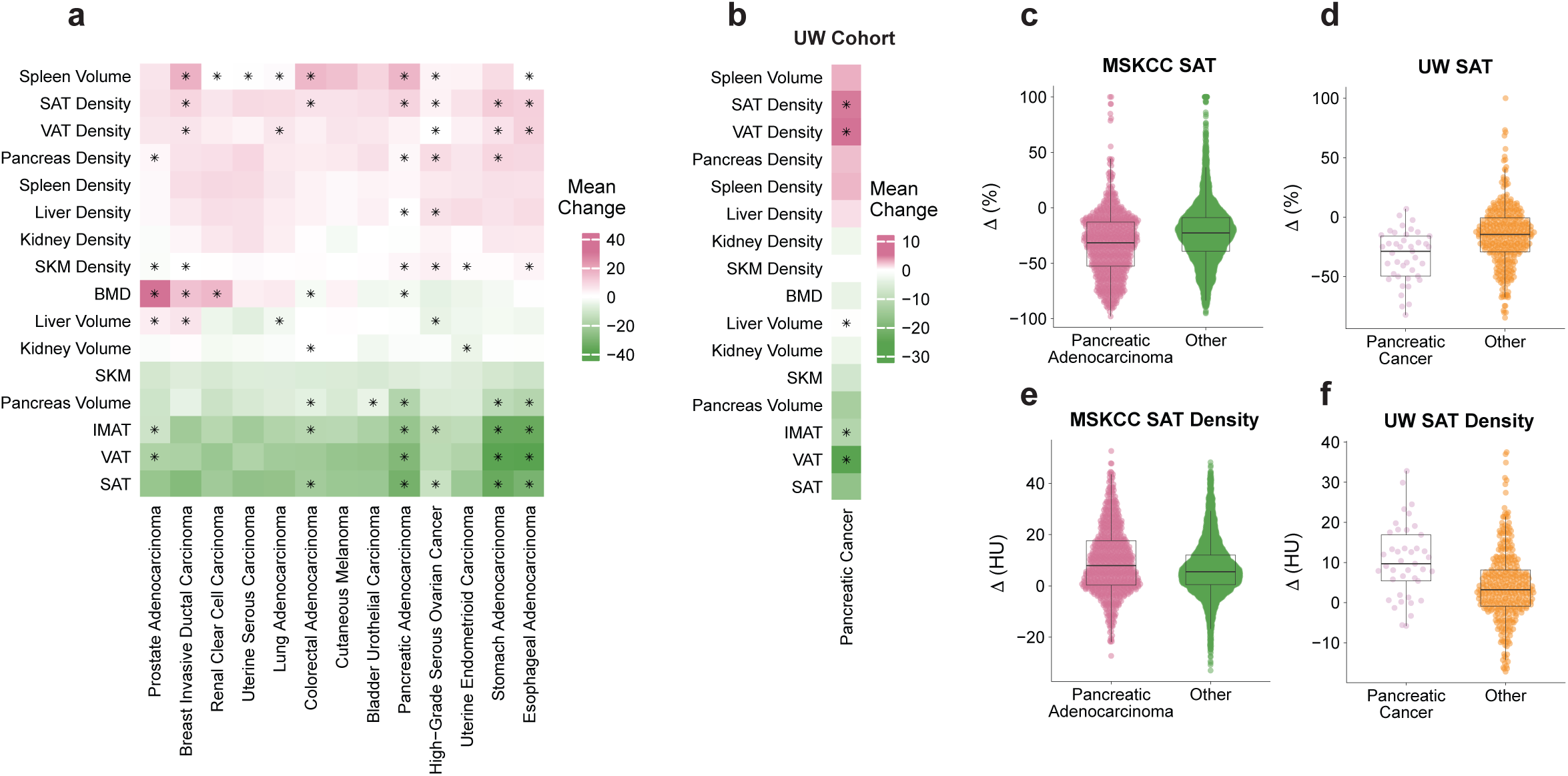
Body composition changes during cachexia vary by cancer type. **a.** Heatmap representing the mean change within-cancer type body composition change from MSKCC primary cohort. “*” designates q < 0.05 from multivariate glm comparing each cancer type versus the rest. **b.** Comparison of body composition changes in pancreatic cancer versus rest in UW validation cohort with univariate glm. “*” designates q < 0.05. **c-d.** SAT changes in pancreatic versus other cancer types in MSKCC, and UW cohorts, respectively. Boxplots indicate mean and interquartile range; error bars, s.e.m. **e-f.** Same as in c-d., but for SAT density.

### Discovery and features of subtypes of cachexia reflecting anatomic evolution

We hypothesized that heterogeneity in anatomic evolution during cachexia could be the consequence of fundamentally different subtypes of cachexia. To test this in an unbiased manner, we applied k-means clustering to standardized changes in body composition across all segmented tissues and organs (see **Methods; Figure 3a**). We determined k = 3 clusters (herein referred to as Type A, Type B, and Type C cachexia) to be optimal by inspecting the elbow in the within-cluster sum-of-squares curve and confirming that additional clusters provided limited improvement and reduced clinical interpretability (**Supplementary Figure 3a**).

**Figure 3:**
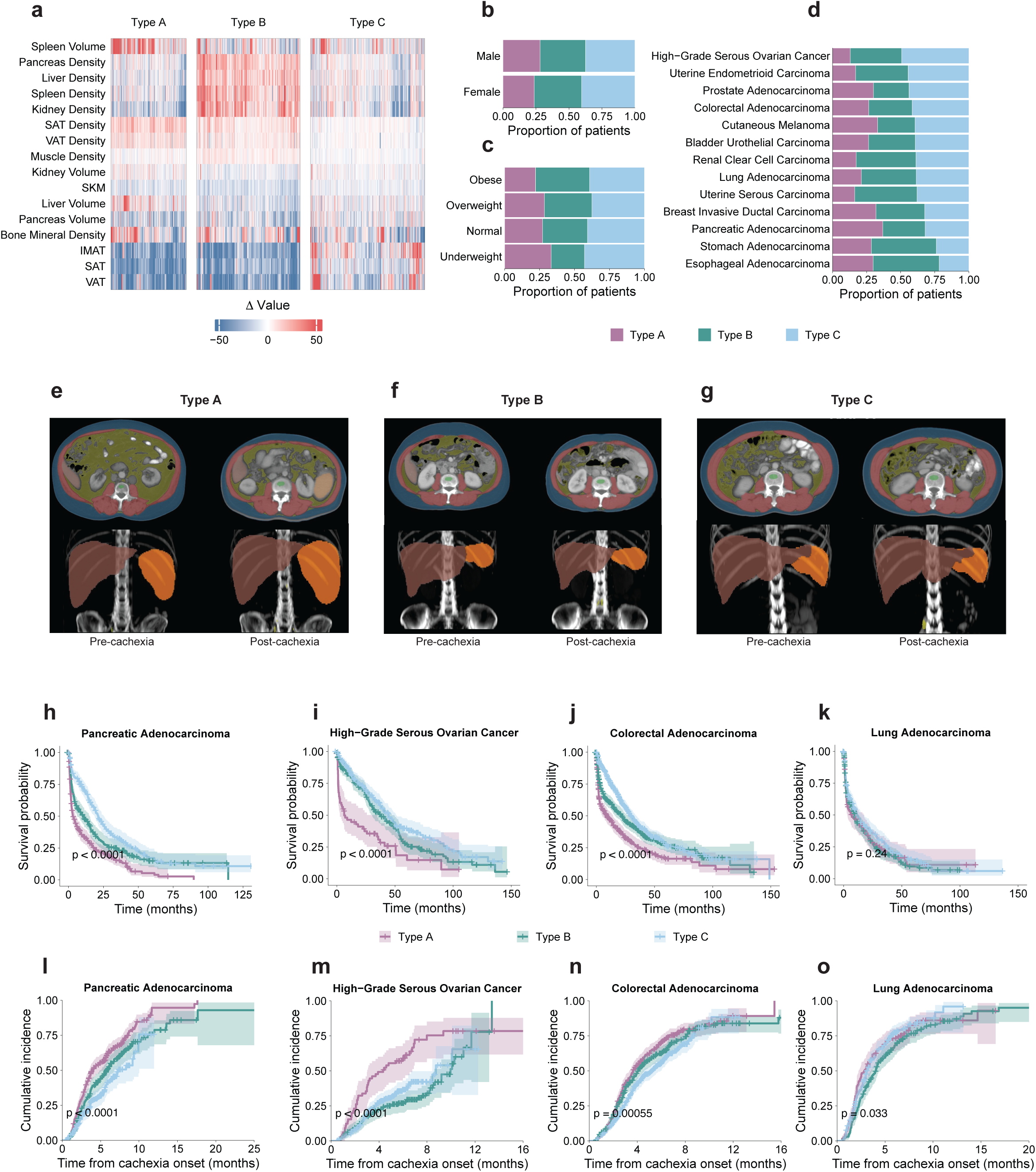
Subtyping cancer cachexia. **a.** Heatmap of body composition changes, separated by k-means derived clusters. Values are clipped at -50 and 50 for visualization. **b.** Sex distribution across clusters. **c.** BMI class at start of cachexia distribution across clusters, **d.** Cancer type distribution across clusters. **e.** Representative OSCAR segmentation example of a Type A male colorectal adenocarcinoma patient, with -28% SAT, -40% VAT, -14% SKM, +10% liver volume, +76% spleen volume. **f.** Representative Type B female pancreatic adenocarcinoma patient, which had a -45% change in SAT, -53% in VAT, -9% SKM, -27% in liver volume, and -13% in spleen volume. **g.** Representative Type C female lung adenocarcinoma patients, with -10% SAT, -25% VAT, -11% SKM, -6% liver volume, -15% spleen volume. **h-k.** Kaplan-Meier curves modelling overall survival from end of cachexia across clusters. P-values from log-rank test. **l-o.** Cumulative incidence for first progression event from cachexia onset. P-values from log-rank test.

The magnitude and compartment-specific nature of weight loss varied significantly across subtypes. BMI loss was substantially greater in Type A and B, where 57.5% and 58.7% of patients, respectively, lost more than 10% of their BMI, compared with only 25.4% of Type C cachexia (**Supplementary Figure 3b**). We observed a modest enrichment for males in Type A cachexia (Chi-Squared P = 0.00251) (**Figure 3b**), and a BMI-dependent association with cluster membership, such that patients with lower pre-cachexia BMI were more likely to belong to Type A (Chi-Squared P = 0.00659) (**Figure 3c**). Consistent with our prior analysis of cancer type specific body composition changes, we identified 8/39 significant association between cancer types and cachexia subtype (Benjamin-Hochberg-corrected post-hoc chi-squared **Supplementary Table 4**; **Figure 3d**), including enrichment for pancreatic adenocarcinoma in Type A cachexia (Chi-Squared q = 1.26×10^-11^, OR = 1.89), stomach adenocarcinoma for Type B cachexia (Chi-Squared q = 0.0271, OR = 1.71), and high-grade serous ovarian cancer for Type C cachexia (Chi-Squared q = 4.36×10^-5^, OR = 1.60).

We examined in detail the specific anatomical changes defining each cachexia subtype by performing pairwise comparisons between cachectic types (**Figure 3e-g**). Key body composition changes across the period of cachectic weight loss were notably more depleted in Types A and B relative to Type C, with significant SAT loss (Type A Welch two sample t-test p < 2.2×10^-16^, Cohen’s d = -1.60; Type B Welch two sample t-test p < 2.2×10^-16^, Cohen’s d = -1.09), VAT loss (Type A Welch two sample t-test p < 2.2×10^-16^, Cohen’s d = -1.05; Type B Welch two sample t-test p < 2.2×10^-16^, Cohen’s d = -1.08), IMAT loss (Type A Welch two sample t-test p < 2.2×10^-16^, Cohen’s d= -1.48; Type B Welch two sample t-test p < 2.2×10-^16^, d = -1.51), and SKM loss (Type A Welch two sample t-test p < 2.2×10^-16^, Cohen’s d= -0.481; Type B Welch two sample t-test p < 2.2×10^-16^, d = -0.756), relative to Type C (**Figure 4a-c**). Muscle density changes were elevated in both Type A and B (Type A Welch two sample t-test p = 9.4×10^-14^, Cohen’s d = 0.284; Type B Welch two sample t-test p < 2.2×10^-16^, Cohen’s d = 0.989), relative to Type C (**Figure 4d**). Interestingly, we noted that within Type A and B, loss of SAT, VAT, and SKM were linearly associated across tissues (**Supplementary Figure 3c-d**), an effect not strongly observed in Type C. Thus, coordinated depletion of muscle and adipose is a characteristic of Type A and Type B patients, but not of Type C patients, aligning with previous reports that BMI fails to capture SKM loss^21^ and is not sufficient to establish cachexia^2,22^. Given that wasting adipose and muscle tissue are the distinguishing clinical features of cachexia, we designed subsequent analyses comparing patients of Type A and Type B cachexia (*i.e.* the most profound wasting phenotypes) to patients with Type C cachexia.

**Figure 4:**
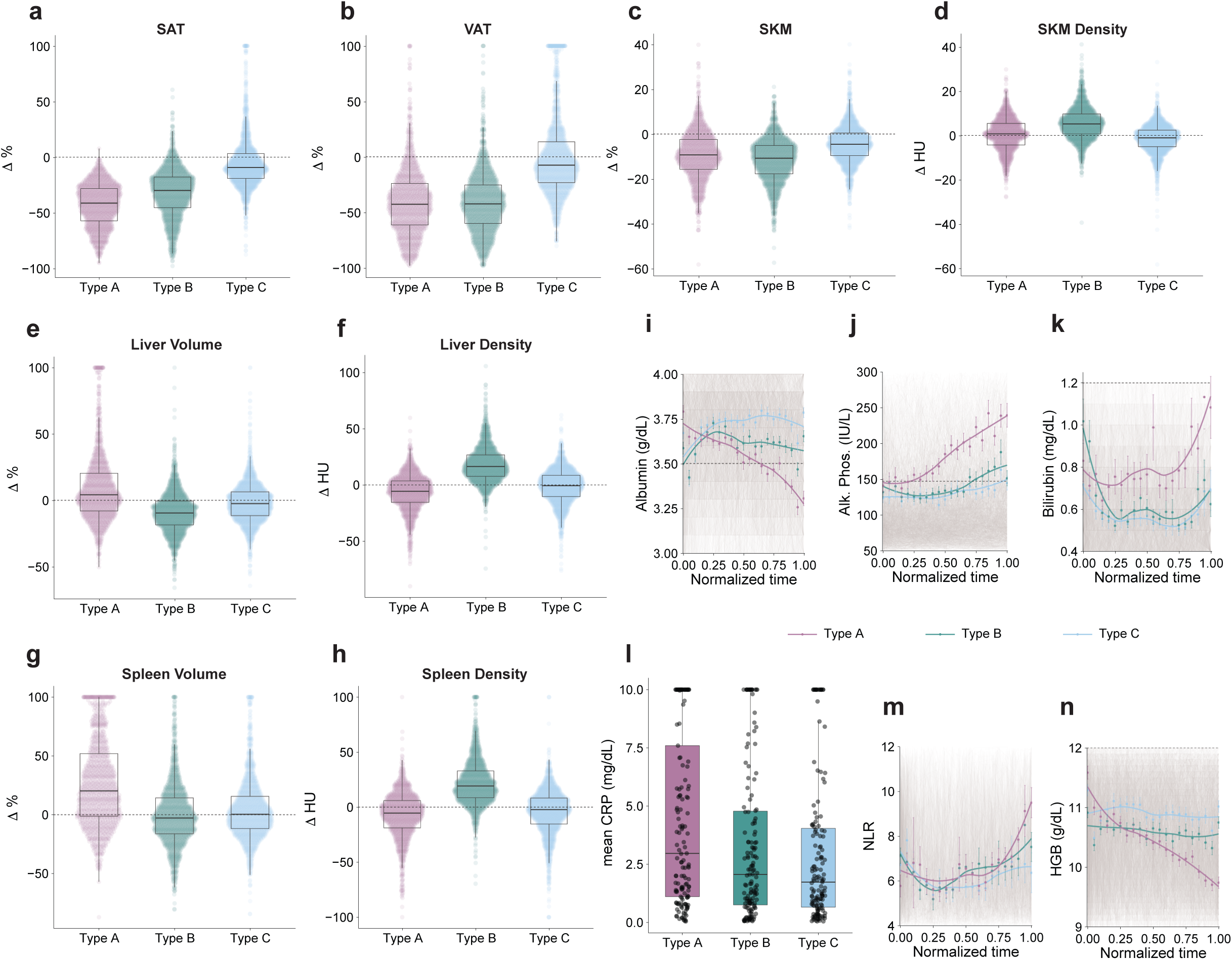
Phenotyping cachectic subtypes. a-h. Pairwise comparisons of key compartment changes across subtypes of cachexia. Boxplots indicate mean and interquartile range; error bars, s.e.m. The dotted line separates gain and loss at zero. **i-k, m-n.** Time normalized blood lab values for albumin, alkaline phosphatase, neutrophil-to-lymphocyte ratio, and hemoglobin. Dots represent binned mean values at 0.05 intervals, line is loess fit. Error bars, 95% confidence intervals. Dotted horizontal lines represent normal laboratory ranges. **l.** Comparison of mean CRP during cachexia across subtypes. Pairwise Welch two sample t-test P = 0.017 (Type A vs. Type C); P = 0.042 (Type A vs, Type B); P = 0.575 (Type B vs. Type C).

Organ-level changes were markedly different across cachexia subtypes. Patients who experienced Type B cachexia exhibited coordinated decreases of all organs volumes, inclusive of the liver (Welch two sample t-test p < 2.2×10^-16^, Cohen’s d = -0.449), the spleen (Welch two sample t-test p = 0.00057, Cohen’s d = -0.148), the pancreas (Welch two sample t-test p < 2.2×10^-16^, Cohen’s d = -0.512), and the kidney (Welch two sample t-test p = 0.049, Cohen’s d = -0.219), and pronounced augmentation in density in liver (Welch two sample t-test p < 2.2×10^-16^, Cohen’s d = 1.22), spleen (Welch two sample t-test p < 2.2×10^-16^, Cohen’s d = 1.19), pancreas (Welch two sample t-test p < 2.2×10^-16^, Cohen’s d =1.41), and kidney (Welch two sample t-test p < 2.2×10^-16^, Cohen’s d = 1.26) (**Figure 4e-h; Supplementary Figure 3e-j**). Intracellular lipid content (steatosis) has characteristically lower CT attenuation values^23,24^, and is a well-established determinant of overall lower tissue density. The synchronous atrophy of organ, adipose, and muscle areas/volumes in combination with increased organ density is therefore suggestive of profound intracellular lipid loss in Type B cachexia, and may represent anorexic wasting.

In contrast to Type B cachexia, patients with Type A cachexia had large, progressive enlargement of splenic volume (Welch two sample t-test p < 2.2×10^-16^, Cohen’s d = 0.682), as well as small progressive enlargement of hepatic volume (Welch two sample t-test p < 2.2×10^-16^, Cohen’s d = 0.512), with concomitant decreases in liver density (Welch two sample t-test p < 2.2×10^-16^, Cohen’s d = -0.323) and in splenic density (Welch two sample t-test p = 0.0016, Cohen’s d = - 0.124). Pancreas volume was reduced (Welch two sample t-test p < 2.2×10^-16^, Cohen’s d = - 0.575), but density (Welch two sample t-test p = 0.1, Cohen’s d = -0.0606), and kidney volume was negligibly enlarged (Welch two sample t-test p = 0.049, Cohen’s d = 0.0679) while kidney density was reduced (Welch two sample t-test p < 2.2×10^-16^, Cohen’s d = -0.470) (**Figure 4e-h; Supplementary Figure 3e-j**). These observations potentially align with murine models of cachexia that have identified liver inflammation as an early event during cachexia evolution^25^ and liver dysfunction as a putative driver of cachexia through impaired hepatic fat metabolism^26,27^.

To assess the robustness of our subtype discovery, we repeated unsupervised subtype analysis in the UW validation cohort. We recovered three clusters with characteristics that aligned with our previously described anatomical subtypes (**Supplementary Figure 4b**). These included two clusters (Type A and Type B) again demonstrating profound SAT loss across weight loss episodes (Type A Welch two sample t-test p < 2.2×10^-16^, Cohen’s d = -1.61; Type B Welch two sample t-test p = 2.7×10^-14^, Cohen’s d = -1.01), VAT loss (Type A Welch two sample t-test p < 2.2×10^-16^, Cohen’s d = -1.34; Type B p < 2.2×10^-16^, Cohen’s d = -1.12), and SKM loss (Type A Welch two sample t-test p = 0.0141, Cohen’s d = -0.327, Type B p = 0.0025, Cohen’s d = -0.409) (**Supplementary Figure 4c-e**). Patients with Type A cachexia in the UW cohort demonstrated (consistent with the MSKCC discovery cohort) progressively increasing liver volume (Welch two sample t-test p = 0.0242, Cohen’s d = 0.220) and increasing spleen volume (Welch two sample t-test p = 0.00024, Cohen’s d = 0.437) and associated decreases in liver density (Welch two sample t-test p = 0.0091, Cohen’s d = -0.296) and spleen density (Welch two sample t-test p = 0.00063, Cohen’s d = -0.430) (**Supplementary Figure 4f-i**). In contrast, patients with Type B cachexia in the UW cohort exhibited atrophy of organ volumes such as the pancreas (Welch two sample t-test p = 2.4×10^-5^, Cohen’s d = -0.0186) and kidney volume (Welch two sample t-test p = 0.00203, Cohen’s d = -0.411), and concomitant increases in organ densities for both the pancreas (Welch two sample t-test p < 2.2×10^-16^, Cohen’s d = 1.39) and the kidney (Welch two sample t-test p < 2.2×10^-16^, Cohen’s d = 1.4) (**Supplementary Figure 4j-m**). Thus, these data establish that cachectic weight-loss episodes can be reproducibly discretized into three broad patterns of anatomical evolution across multiple cohorts of cancer patients.

### Patients with Type A and Type B cachexia have inferior clinical outcomes

Cachexia has been demonstrated to be detrimental to patient health, leading us to hypothesize that patterns of anatomical evolution would correlate with clinical outcomes. To test this, we modeled overall survival (subsequent to the post-cachexia CT scan) in each cancer type as a function of cachexia subtype. Across most cancer types, including pancreatic adenocarcinoma (log-rank P < 0.0001), high-grade serous ovarian cancer (log-rank P < 0.0001), and colorectal adenocarcinoma (log-rank P < 0.0001), both Type A and Type B patients had inferior overall survival compared to Type C, with Type A patients consistently having the more extreme effect (**Figure 3h-j, Supplementary Figure 5a-i**). Lung adenocarcinoma was the exception, where survival probability did not stratify by cachectic subtype (log-rank P = 0.24) (**Figure 3k**).

We previously reported that disease progression is elevated during cachectic episodes^18^. We therefore asked whether cachectic subtypes demonstrate differential rates of disease progression, as assessed through previously reported analysis of radiographic notes from the MSK-CHORD dataset^28^. To do so, we modeled the time to the first disease progression event during weight-loss episodes. Patients with Type A cachexia had higher incidence of progression in pancreatic adenocarcinoma (log-rank P < 0.0001), high-grade serous ovarian cancer (log-rank P = 0.0032), uterine serous carcinoma (log-rank P = 0.0021), and in stomach adenocarcinoma (log-rank P = 0.043) (**Figure 3l-o, Supplementary Figure 5j-r**). Together, these data demonstrate that anatomical subtypes of cachexia are associated with distinct patterns of disease progression.

### Systemic inflammation is a defining feature of Type A cachexia

We reasoned that blood laboratory measurements, in combination with several derived markers such as prognostic nutritional index (PNI) and neutrophil to lymphocyte ratio (NLR) (see **Methods**), could inform our understanding of the basis of dynamic anatomical subtypes of cachexia. We therefore fit linear mixed effects models to assess the serological profiles of Type A and Type B cachexia in comparison to Type C cachexia while controlling for sex, age, BMI, stage, and cancer (**Supplementary Figure 6a; Supplementary Table 5**). Patients with either Type A or Type B cachexia respectively presented with patterns of malnutrition and anemia, marked by low albumin (Type A log_2_(fold-change) = -0.032, q = 4.67×10^-9^; Type B log_2_(fold-change) = -0.0194 q =1.20×10^-4^), low hemoglobin (HGB) (Type A log_2_(fold-change) = -0.0445, q = 2.63×10^-12^ & log_2_(fold-change) = -0.0238, q = 5.14×10^-5^), low hematocrit (HCT) (Type A log_2_(fold-change) = -0.0407, q = 1.56×10^-12^; Type B log_2_(fold-change) = -0.0216, q = 5.03×10^-5^), and low red blood cell count (RBC) (Type A log_2_(fold-change) = -0.0444, q = 3.26×10^-12^; Type B log_2_(fold-change) = -0.0272, q = 4.06×10^-6^), as has been previously described to occur in cachexia^29–31^.

Several lab values distinguished Type A from Type B cachexia. Patients with Type A cachexia presented with elevated markers of systemic inflammation, such as high neutrophil-to-lymphocyte ratio (NLR) (log_2_(fold-change) = 0.0865, q = 3.20×10^-3^), and of liver abnormalities, such as high alkaline phosphatase (log_2_(fold-change) = 0.180, q = 1.38×10^-13^), high aspartate aminotransferase (AST) (log_2_(fold-change) = 0.108, q = 1.08×10^-7^), and high bilirubin (log_2_(fold-change) = 0.121, q = 7.28×10^-8^), whereas only patients with Type B cachexia had low alanine aminotransferase (ALT) levels (log_2_(fold-change) = -0.0755, q = 1.34×10^-3^). Consistent with a pattern of elevated inflammation in Type A cachexia, in a subset of n = 1,627 patients where additional blood laboratory panels were ordered, patients with Type A cachexia demonstrated elevated C-reactive protein (log_2_(fold-change) = 0.511, q = 0.014) and ferritin (log_2_(fold-change) = 0.520, q = 7.85×10^-5^), which are acute phase reactants and widely used clinical markers of systemic inflammation^32^ (**Figure 4l; Supplementary Figure 6b; Supplementary Table 6**). Liver and bone metastasis can produce elevated levels of alkaline phosphatase and other liver function tests, potentially confounding the serologic signatures of cachectic subtypes. However, *post-hoc* re-analysis of n=1,531 with no clinical history of liver or bone metastasis confirmed a depletion of albumin in both Type A (log_2_(fold-change) = -0.0385, P = 5.69×10^-5^) and Type B cachexia (log_2_(fold-change) = -0.0233, P = 1.84×10^-3^), and elevation of Alk. phos. (log_2_(fold-change) = 0.0695, P = 0.0238), and AST (log_2_(fold-change) = 0.108, P = 2.74×10^-4^) in patients with Type A cachexia, confirming that serologic signatures of cachectic subtypes are independent of the presence of liver metastasis.

We next examined how key serological markers change dynamically across cachectic episodes (**Figure 4i-k; m-n**). We aggregated laboratory values into 20 time-normalized bins and qualitatively compared subtype trajectories during cachexia. To assess statistical differences in the blood laboratory trajectories, we conducted pairwise comparisons between cachectic subtypes, focusing on changes between the end to the start of cachexia, and identified 25 FDR significant changes in Type A relative to Type C, and 10 in Type B relative to Type C. In agreement with our previous results, the progression of inflammation and abnormalities on liver function tests was significantly more pronounced in Type A patients compared to both Type B and Type C patients. Albumin and HGB, both negative acute phase reactants to inflammation, acutely drop in Type A (Albumin Welch two sample t-test q = 1.47×10^-47^, log_2_(FC) = -3.42; HGB Welch two sample t-test q = 9.49×10^-35^, log_2_(FC) = -1.43), and are mildly depressed in Type B (Albumin two sample t-test q = 0.00603, log_2_(FC) = -1.47; HGB two sample t-test q = 0.499, FC = 1.13) patients. NLR and Alk. phos. increase strongly at the end of cachexia Type A (NLR Welch two sample t-test q = 3.07×10^-8^, log_2_(FC) = -3.90; Alk. phos. Welch two sample t-test q = 8.90×10^-12^, log_2_(FC) = - 2.09;) but not in Type B (NLR Welch two sample t-test q = 0.629, log_2_(FC) = -1.22; Alk. phos. Welch two sample t-test q = 0.908, log_2_(FC) = 0.096). These observations demonstrate how serology dynamically tracks with changes to patient anatomy, and indicate that systemic inflammation is a hallmark of Type A cachexia.

### Molecular phenotypes of primary tumors in patients experiencing cachectic wasting

Given that cachexia occurs secondary to cancer, it has been suggested that the tumor is a key driver of the disease^2,26,33^. However, testing if subtypes of cachexia are associated with differential molecular phenotypes in tumors is challenging, as it requires (1) integrated collection of body weight, radiographic, and molecular data on cancer patients to call cachectic episodes, and (2) collection and profiling of tumor tissue at a consistent time-point (*i.e.* during cachexia) to ascertain phenotypic associations. We identified an institutional dataset of single-cell RNA-sequencing from pancreatic adenocarcinoma patients in arm A of the POLAR trial^34^, which evaluated maintenance pembrolizumab plus olaparaib following platinum-based chemotherapy in *BRCA1*/*BRCA2*–mutated or *PALB2*-mutated HR-deficient patients. We identified five patients from this trial for whom we had matched body weight data (and could confirm that patients experienced a cachectic episode), pre- and post-cachexia contrast-enhanced CT scans, and associated single-cell RNA sequencing data from tumor tissue collected during the cachectic weight loss time period^35^. We mapped patients onto anatomical subtypes of cachexia using our CT-derived body composition approach, identifying 3 Type A and 2 Type C patients (with no Type B patients likely due to small sample size) (**Figure 5a**).

**Figure 5:**
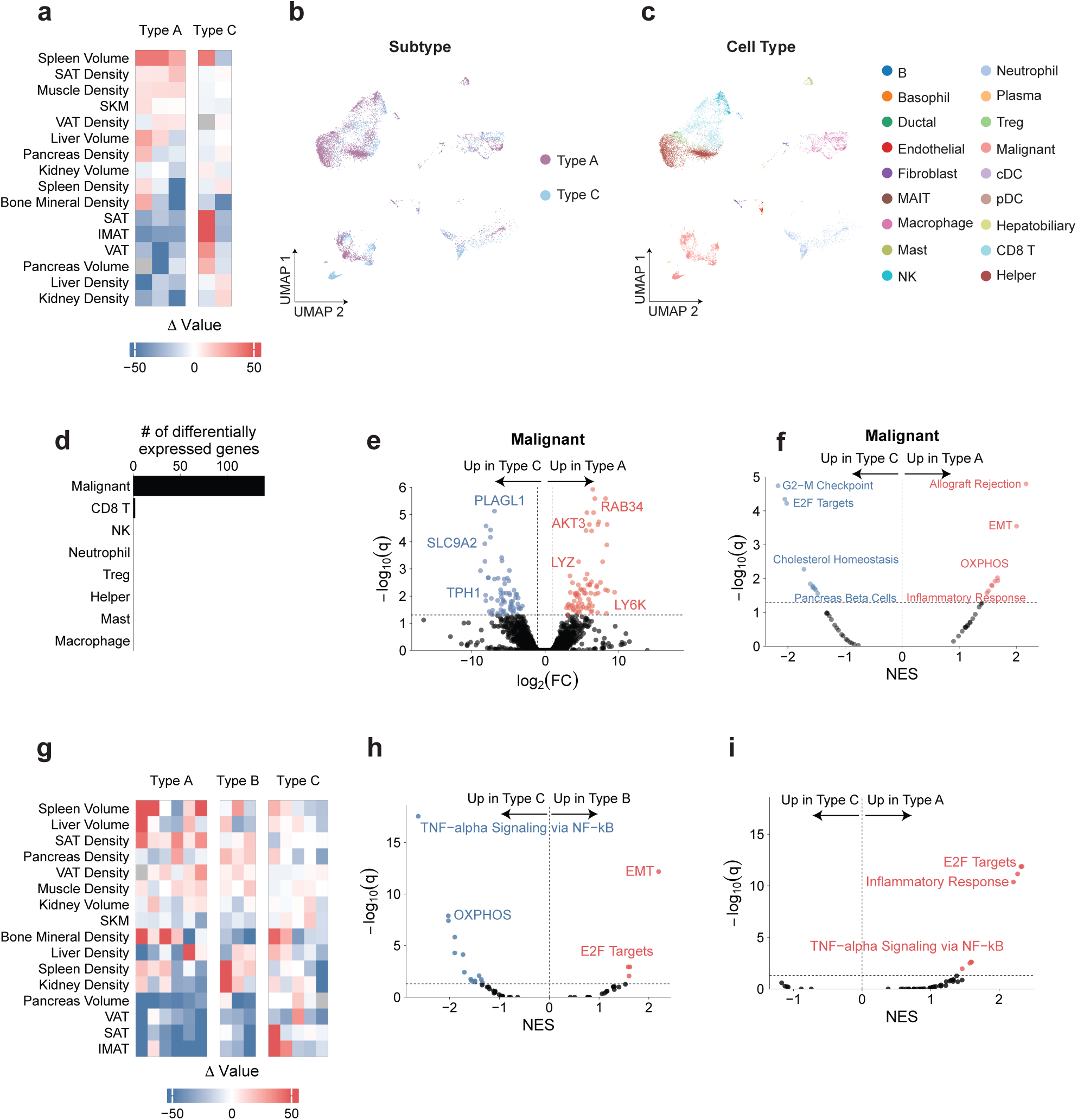
Molecular features of the cachectic subtypes. **a.** Heatmap of body composition changes by subtypes in PDAC tumor cohort. **b-c.** UMAP plot of cell profiled by scRNA sequencing colored by cachectic subtype (a), and cell type (b). **d.** Number of significantly differentially expressed genes in each cell type. **e.** Differential gene expression results in malignant cells, comparing Type A to type A. **f.** GSEA results in malignant cells comparing Type A to Type C. **g.** Heatmap of body composition changes by subtypes in PDAC liver biopsy cohort. **h-i.** GSEA results from bulk RNA-sequencing of pre-metastatic liver samples comparing Type B to Type C and Type A to Type C.

We hypothesized that tumors collected from patients with different subtypes of cachexia would potentially demonstrate different gene expression patterns. We quantified gene expression profiles across 18 distinct cell types (**Figure 5b-c**). On a per-cell-type basis, we compared (pseudobulked) gene expression profiles between Type A and Type C patients. We observed that gene expression differences between tumors of Type A and Type C cachexia were primarily concentrated in malignant cells. Specifically, in the malignant cell population, we identified 75 significantly upregulated and 65 significantly downregulated differentially expressed genes between Type A and Type C patients (**Figure 5d-e**). In contrast, across all other cell types, we only identified 2 significantly differentially expressed genes in CD8+ T cells, suggesting that in these data, molecular differences between cachectic subtypes are primarily driven by tumor cells (**Figure 5d)**. The concentration of gene expression changes in malignant cells may be potentially ascribable to low cell counts in non-immune populations, although we observed comparable numbers of T cells and malignant cells but very few differentially expressed genes in the T cell clusters.

To understand whether these gene expression differences were enriched in specific pathways, we carried out gene-set enrichment analysis (GSEA). Focusing initially on the malignant cells, we identified 19 gene sets with statistically significant changes in Type A relative to Type C cachexia, most notably upregulation of inflammatory response (NES = 1.51, q = 0.023) and downregulation of cholesterol homeostasis (NES = -1.57, q = 0.0183) (**Figure 5f**). These findings indicate that the tumor cells from patients with Type A cachexia demonstrate signatures of increased inflammatory signaling. We secondarily considered that, despite a lack of differential gene expression on a single gene basis, subtle differences in gene expression at the pathway level may be evident in other cell types. We therefore carried out GSEA across other cell types, again comparing patients with Type A to patients with Type C cachexia. We identified a total of 25 significantly upregulated and 88 significantly downregulated pathways in Type A compared to Type C across all immune cell-types (**Supplementary Table 7**). Inflammatory response was significantly upregulated in macrophages (NES = 1.89, q = 0.0016) but significantly downregulated in CD8+ T cells (NES = -1.95, q = 0.000756) and in NK cells (NES = -1.76, q = 0.000155). We identified additional evidence of inflammation in Type A through upregulation of TNF-alpha signaling via NF-kB in both macrophages (NES = 2.53, q = 0.00) and mast cells (NES = 1.49, q = 0.00349). These data indicate that tumor cells in Type A cachexia have gene expression patterns clearly consistent with elevated inflammatory signaling, whereas other cell types in the PDAC tumor microenvironment have subtle and more heterogeneous patterns of inflammatory gene expression.

### Molecular signatures of systemic inflammation in organ tissue from Type A cachexia patients

The findings in **Figure 4** and **Figure 5a-f** suggest that systemic inflammatory signaling is highly active in Type A cachexia. Such inflammatory signaling has been previously described in murine models of cancer cachexia^26,36,37^, and would likely provoke cellular responses in both tumor tissue and uninvolved organs throughout the body. In particular, given the pattern of progressive liver volume increase, liver density decrease, and elevation of liver function tests in patients with Type A cachexia, we hypothesized that otherwise uninvolved liver (*i.e.* without any evidence of malignant involvement) from patients with Type A cachexia would demonstrate evidence of elevated inflammatory signaling. Although molecular measurements of non-malignant tissue (and in particular, measurements of uninvolved organs such as the liver) are rare, a recent study from our institution described the transcriptomic profiling of non-malignant liver in n=44 patients undergoing surgical procedures for pancreatic cancer. Of these, 14 patients had longitudinal BMI data, demonstrated weight loss consistent with cachexia, and had eligible abdominal CT scans flanking the pre- and post-cachexia time points (see **Methods**). Classification based on anatomical changes across cachexia ultimately assigned 6 patients to Type A cachexia, 3 patients to Type B cachexia, and 5 patients to Type C cachexia (**Figure 5g**).

To determine whether patients with distinct subtypes of cachexia demonstrated differential molecular phenotypes in the uninvolved, pre-cachectic liver, we carried out differential gene expression analysis, and applied gene set enrichment analysis to these findings using MSigDB Hallmark genesets (**Supplementary Table 8**). This approach identified that patients with Type B cachexia demonstrated upregulation of epithelial mesenchymal transition (NES = 2.20, q = 6.69×10^-13^) and E2F targets (NES = 1.59, q = 1.17×10^-3^), relative to patients with Type C cachexia. In contrast, patients with Type A cachexia demonstrated upregulation of inflammatory response (NES = 2.21, q = 4.24×10-^11^) and TNF-alpha signaling via NF-kB (NES = 1.60, q = 2.49×10^-13^), relative to patients with Type C cachexia (**Figure 5h-i**). These findings indicate that Type A cachexia is characterized by systemic inflammation that includes, at minimum, organs without evidence of cancer^38^.

## Discussion

In order to study and ultimately therapeutically intervene in cancer cachexia, we must look at patients. The premise of this work is that cross-sectional radiographic data longitudinally collected as part of clinical care can be used to watch the cachectic process unfold. This study applies computational methods to extract interpretable information on the quality and quantity of muscle, fat, and organs from each CT scan, enabling an evolutionary approach to interrogating cachexia primarily through the lens of patient anatomy.

Among our several discoveries, two are especially fundamental and build on a rich prior literature in cachexia in model systems and human patients. The first key discovery is that, by systematically measuring anatomical evolution across thousands of patients, we determined that anatomical changes beyond the muscle and fat are key elements of anatomical remodeling during cachexia. Although the most profound and pervasive changes are in adipose tissue (**Figure 1**), it is instead changes to other organs that discriminate between the two most aggressive subtypes of cachexia (**Figure 3**).

The second key discovery is that clinically meaningful subtypes of cachexia can be defined based on anatomical evolution rather than molecular profiling alone. We describe three subtypes of cachexia, including an inflammatory-like Type A, an organ-atrophy-like Type B, and a mild Type C, based exclusively on evolutionary changes in anatomy. This approach stands in contrast to the dominant paradigm in cancer biology, where subtypes are typically derived from genomic or transcriptomic features within tumors. These differences could inform stratified clinical trials in which therapies are matched to the dominant biological mechanism of each subtype—an approach that has been transformative in oncology but has yet to be applied to cachexia. Furthermore, the ability to identify these subtypes non-invasively through CT-based anatomical phenotyping opens a realistic path to clinical implementation. In this framework, one can envision a future in which therapeutic strategies for cachexia are tailored not to a one-size-fits-all syndrome, but to biologically coherent, anatomically defined patient states.

More broadly, this work provides a framework for linking macroscopic physiological remodeling with microscopic molecular programs. The anatomical features defining Type A cachexia, most notably progressive hepatosplenic enlargement, are accompanied by systemic inflammatory signatures in peripheral blood and are recapitulated at the transcriptional level in primary tumor and uninvolved liver tissue, where pathways such as TNFα signaling via NF-κB and inflammatory response are upregulated. These findings support a model in which systemic inflammation is not confined to circulating factors or tumor-adjacent microenvironments, but is reflected in, and potentially propagated through, organ-level dysfunction in metabolically central tissues. Single-cell analyses of primary tumors reveal that these anatomical subtypes are associated with cell-type-restricted transcriptional phenotypes, including differential activation of inflammatory signaling across tumor and immune compartments. Together, these observations suggest that anatomical subtypes of cachexia represent emergent, organism-level states that are mirrored across scales, from organ physiology to cell-intrinsic transcriptional programs. In this sense, cachexia can be understood not only as a syndrome of wasting, but as a coordinated reprogramming of the human body that bridges gross physiology with molecular biology.

### Limitations

Several limitations of this work should be acknowledged. First, our study is based on a retrospective cohort from a single referral center, and some patients may have already experienced significant wasting prior to presentation, potentially underestimating the full spectrum of anatomical changes. Second, we cannot definitively establish the causal direction between hepatosplenic changes and systemic wasting—whether changes to these organs drive cachexia, respond to it, or both remains an open question. Finally, the transcriptomic analyses of liver tissue and tumor tissue were performed on a small number of patients, limiting statistical power and precluding the detection of more subtle molecular differences. Future studies incorporating larger, multi-institutional cohorts across diverse malignancies will be needed to establish the generalizability of these subtypes and their associated hepatic molecular programs.

## Data availability

Gene expression data for PDAC liver biopsies can be accessed as GSE245535^39^ (bulk mRNA-seq). Single cell RNA sequencing data for POLAR trial patients can be requested from the investigators of the POLAR trial upon reasonable request.

## Code availability

Access to the OSCAR toolkit is available upon request. Scripts used to process segmentation outputs, perform subtype discovery, and produce final figures are available at https://github.com/reznik-lab/cachexia_subtypes_bodycomp.

## Acknowledgements

We would like to thank all the members of the Reznik laboratory for exciting and productive discussion. We would also like to acknowledge very useful feedback from the CANCAN Cancer

Grand Challenges Team. This work was supported by NIH R37 CA276200, NCI P30 CA008748, NIH OT2 CA278609, Pershing Square Sohn Cancer Research Foundation. This research was also supported by the Specialized Program of Research Excellence (SPORE) program, through the National Cancer Institute (NCI), grant P50CA269011. The content is solely the responsibility of the authors and does not necessarily represent the official views of the NIH. This work was also supported by the Cancer Grand Challenges partnership funded by Cancer Research UK (CGCATF2021/100022) and the NCI (CA278685). R.R.K. is supported, in part, by a Department of Defense Early Career Investigator grant (KCRP-AKCI, W81XWH-21-1-0942) and a Focus Award in partnership with the Kidney Cancer Association and Joey’s Wings Foundation. This work was also supported by NIH/NCI SPORE Pancreas (P50 CA257881-01A1), Paul Calabresi Career Development Award for Clinical Oncology (K12 CA184746), Cancer Center Support Grant/Core Grant (P30 CA008748), Break Through Cancer, Parker Institute for Cancer Immunotherapy, The Society of MSK.

## Methods

### Patient cohort and cachexia identification

The cohort was derived from MSK-IMPACT, a prospective observational study of tumor evolution at Memorial Sloan Kettering Cancer Center. We obtained multimodal radiologic, clinical, and serologic data on 79,240 patients consented to the MSK-IMPACT prospective clinical sequencing protocol as part of their routine clinical care. The study was approved by the MSKCC Institutional Review Board (IRB), and all patients provided written informed consent for review of medical records. Retrospective BMI measurements were available for 59,493 patients, where we determined which patients over the age of 18 experienced greater than 5% weight loss within 6 months or 2% weight loss within 6 months for patients with BMI < 20^18^. To study changes in body composition across these cachectic windows, we identified the abominable contrast-enhanced CT scan that was obtained within a 90-day window of pre-cachexia and of post-cachexia to obtain a cohort of 4,516 total patients. If a patient experienced more than one cachectic episode, only the first episode was considered. Importantly, for the remainder of our analyses, cachexia start and end dates were considered to be at the time the CT scans were conducted to ensure consistency.

### University of Wisconsin Validation cohort

A separate validation cohort from the University of Wisconsin-Madison (UW Madison) was collected (IRB 2021-1403). This Health Insurance Portability and Accountability Act-compliant retrospective cohort study was approved by the institutional review boards of UW Madison. The requirement for informed consent was waived. All adult patients over the age of 18 with a CT of the abdomen between January 2001 and October 2023 were identified. From this group, patients with a known cancer diagnosis, sufficient BMI measurements to identify cachexia events, and appropriate timing of the CT scans (within 90 days of identified cachexia events) were retained, yielding a total cohort of 392 patients.

### Automatic segmentation of body composition changes

Each CT scan was processed using an automated CT body composition pipeline consisting of standardized preprocessing, anatomical localization, multi-organ/tissue segmentation, and quantitative biomarker extraction^14^. Input DICOM images were converted to NIfTI format, reoriented to a consistent anatomical coordinate system, and resampled to a uniform voxel spacing. Image intensities were normalized to ensure compatibility across acquisitions. A convolutional neural network–based anatomical localization model was then applied to estimate vertebral landmarks (e.g., T10–L5), enabling consistent identification of anatomical levels for downstream analysis.

Segmentation was subsequently performed using deep learning models based on 3D encoder–decoder architectures (U-Net–like convolutional neural networks), with incorporation of transformer-based components for select whole-body segmentation tasks. These models were used to delineate skeletal muscle, visceral and subcutaneous adipose tissue, and major abdominal organs. From the resulting segmentations, quantitative biomarkers were computed, including cross-sectional area, volumetric measures, and attenuation-based metrics (mean Hounsfield units) at standardized anatomical levels and across volumetric regions. The pipeline was executed within a containerized environment, and summary outputs and segmentation overlays were generated for downstream analysis.

After segmentation, we excluded patients with a calculated total fat area of less than 35 cm² at any time point, as these low values typically reflected segmentation failure in the setting of diffuse anasarca, which obscured boundaries between tissue compartments. We additionally filtered out any values that were outside the physiological range^14^.

### Manual segmentation of body composition

To assess the robustness of the OSCAR method in our cohort, we performed manual SAT, VAT, and SKM segmentation in a subset of 100 CRC patients. Adipose and skeletal muscle composition were analyzed on CT imaging using *TeraRecon Aquarius iNtuition* (TeraRecon Inc.). For each patient, an abdominal series was identified and one axial image at the L3 vertebral level, with slice level confirmation by a radiologist. Adipose tissue compartments were quantified using the built-in fat analysis tool, which automatically segments and reports both SAT and VAT, with manual correction for misclassified regions and boundary discontinuities when present. SKM was quantified on the same slice by manual contouring with the ROI/contour tool to trace the muscle boundary while excluding non-target structures (vertebral bone, intraluminal contents, and imaging artifacts). The same segmentation workflow was applied across available scans, and a subset underwent secondary review to confirm consistency of slice selection and compartment boundaries.

### Calculation of body composition changes

We assessed body composition changes as percent changes for SKM area, SAT area, VAT area, and pancreas, liver, spleen, and kidney volumes.

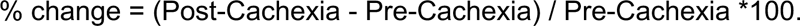

For all densities, including all aforementioned tissues and organs, as well as bone mineral density (all measured in Hounsfield Units), we calculated raw changes, as they are more clinically meaningful given that zero HU is defined as water density and not a physiological setpoint.

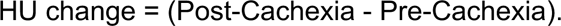

In addition, we computed percent changes for VAT, SAT, and SKM areas at axial levels L1, T10, T12 to ensure robustness of OSCAR segmentation across spinal levels. Finally, we filtered out any patients that had abnormally large changes in SAT or VAT measurements, which we defined as >500%.

### Assessment of normal fluctuations

We assembled a cohort of 280 non-cachectic controls across pancreatic adenocarcinoma, uterine endometrioid carcinoma, lung adenocarcinoma, and renal clear cell carcinoma. To ensure comprehensive coverage over the entire disease trajectory, we selected CT scans to be between three and six months apart, with a minimum of three months and maximum five years total follow-up. We computed normal fluctuations within each organ or tissue by calculating time-normalized percent changes across each consecutive CT scan and taking the per patient median.

### Association of body composition changes across cancer types

To evaluate whether a specific cancer type exhibited distinct patterns of body composition changes relative to other cancers, we fitted a series of linear models for each body composition metric, using cancer type (target type vs. all other cancers) as the main predictor, while controlling for sex, age, and BMI at cachexia onset. P-values were adjusted for multiple hypothesis testing using Benjamin-Hochberg correction. We modeled this as

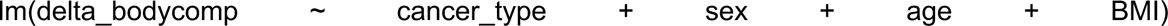

In the UW validation cohort, cancer type annotations were constrained to primary organ site, and demographics were not available. Therefore, we repeated the analysis, restricting only to pancreatic cancer compared to all other cancer types using a univariate linear model.

### Clustering of body composition measurements

Body composition changes were first imputed for missing values using k-nearest neighbors (k = 10) with the VIM:knn() package in R. Imputed values were then z-score transformed across patients to ensure comparable feature contributions. K-means clustering performed on the scaled matrix with k = 3 clusters and nstart = 50 with a set seed. The number of clusters, k, was deduced based on the elbow method, as visualized by the screeplot (**Supplementary Figure 3a**).

### Analysis of serological measurements

We analyzed 30 longitudinal blood lab measurements from the complete blood count panel and the complete metabolic panel. We restricted our analysis to only measurements collected between the pre-cachexia and post-cachexia scans. We included several derived laboratory values to determine inflammation and nutritional status, namely:

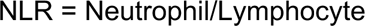

First, we qualitatively assessed differences in key serological trends by calculating the within-cluster mean blood laboratory value at 0.05 normalized time intervals, and plotting 95% confidence intervals. Within cluster trajectories were visualized using loess smoothing from geom_smooth().

To quantitatively evaluate characteristic differences between longitudinal serological values during weight loss we employed a linear mixed effects model, controlling for age at the time of the pre-cachexia scan, sex, stage, BMI, and cancer type as follows. Pairwise comparisons across the clusters were computed using the pairs() function from the emmeans() library.

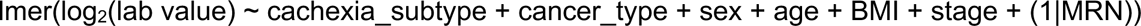

### Overall survival analyses

To examine the clinical profiles of cachectic subtypes, we modeled overall survival within each cancer type using time-to-event analysis, with follow-up time measured from the end of cachexia. Kaplan-Meier curves were used to visualize survival according to subtype, and between-group differences were evaluated with the log-rank test.

### Time to first progression analyses

We modeled the cumulative incidence of the first progression event from cachexia onset as a time-to-event-outcome within each cancer type. As both cachectic start time and progression were radiologically derived, we considered only progression events that occurred at least 15 days after cachexia onset. Differences between cachectic subtypes were assessed using log-rank tests.

### Single-cell analyses in PDAC

We annotated PDAC patients^34^ that went on to develop cachexia by their anatomical subtype. We included only patients that corresponded to arm A of the trial, and had sample collection during their cachectic episode, up to 120 days following the cessation of cachexia.

We proceeded to use PyDEseq2^41^ differential expression analysis on scRNA data in order to identify signatures of cachexia subtypes. Using count data, we created sample-level pseudobulks for each sample/cell-type pair for samples with at least 10 cells of the cell type of interest. Then, we ran PyDEseq2 differential expression contrasting Type A vs. Type C and performed ranked GSEA analysis with gseapy^42^ by ranking genes by Wald statistic and using Hallmark pathways (v2025). P-values were adjusted for multiple hypothesis correction using Benjamin-Hochberg.

### Bulk-liver RNA seq analyses

To determine the molecular phenotype of uninvolved liver tissue across our three subtypes, we examined bulk-liver RNA sequencing data from a subset of pre-metastatic pancreatic adenocarcinoma patients already profiled at our institution^39^. Initially, we had eligible CT scans and successful body composition data for 10/44 patients originally profiled. To boost statistical power, we relaxed the CT scan criteria from 90 days to 270 days from either cachectic start and end dates, and rescued an additional four patients, yielding a final cohort of n=14 with paired bulk-RNA sequencing and radiographically derived cluster annotation.

Differential gene expression analysis was performed using limma v3.60.6 in R (v12.1). The normalized gene expression matrix was log_2_ transformed and the design matrix was constructed to model subtype differences (Type A or Type B) relative to Type C. We applied lmFit() followed by eBayes() with robust and trend estimation enabled. Geneset enrichment analysis was performed on genes ranked by log_2_(fold-change) using Hallmark pathways (v2025). P-values were adjusted for multiple hypothesis correction using Benjamin-Hochberg.

