## Supplemental Figures for "Anatomical dynamics define cancer cachexia subtypes and identify systemic inflammation as a marker of lethal wasting"

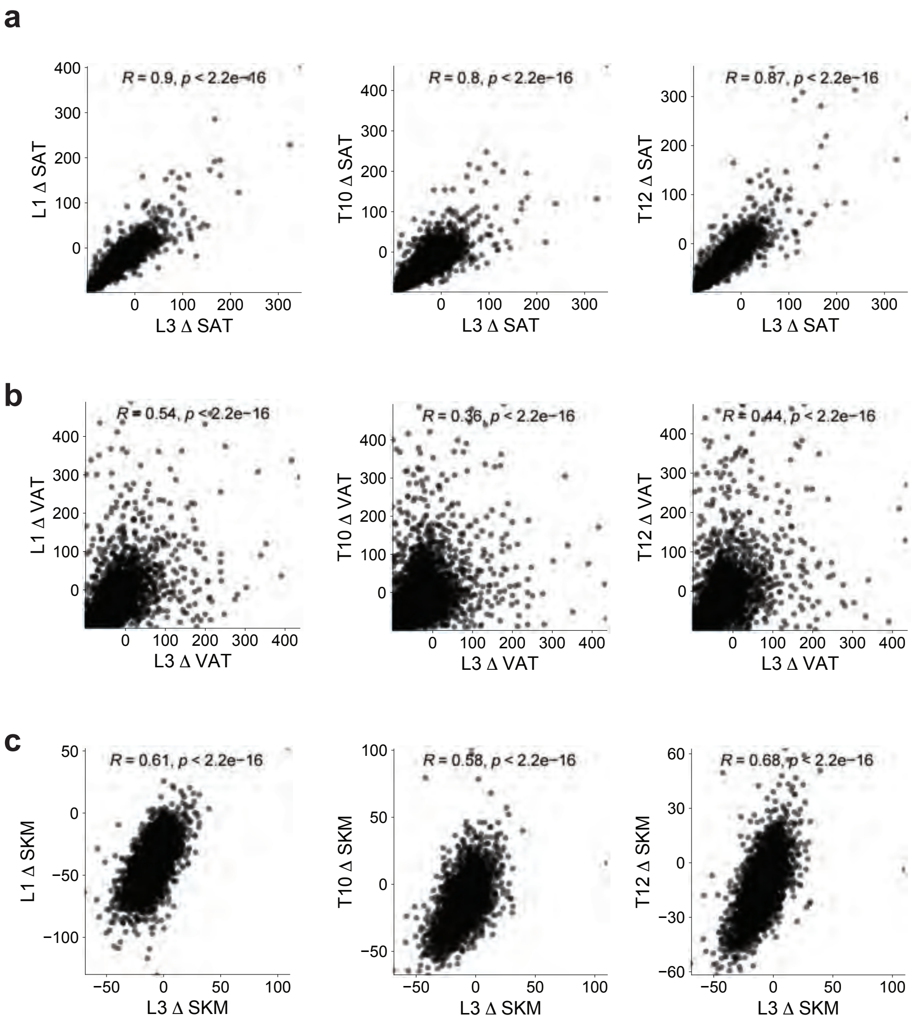


**Supplementary Figure 1: Corroboration of body composition measurements across spinal levels. a-c.** Correlation of body composition changes of L3 against L1, T10, and T12 for SAT, VAT, and SKM. R and P values are derived from Pearson Correlations.

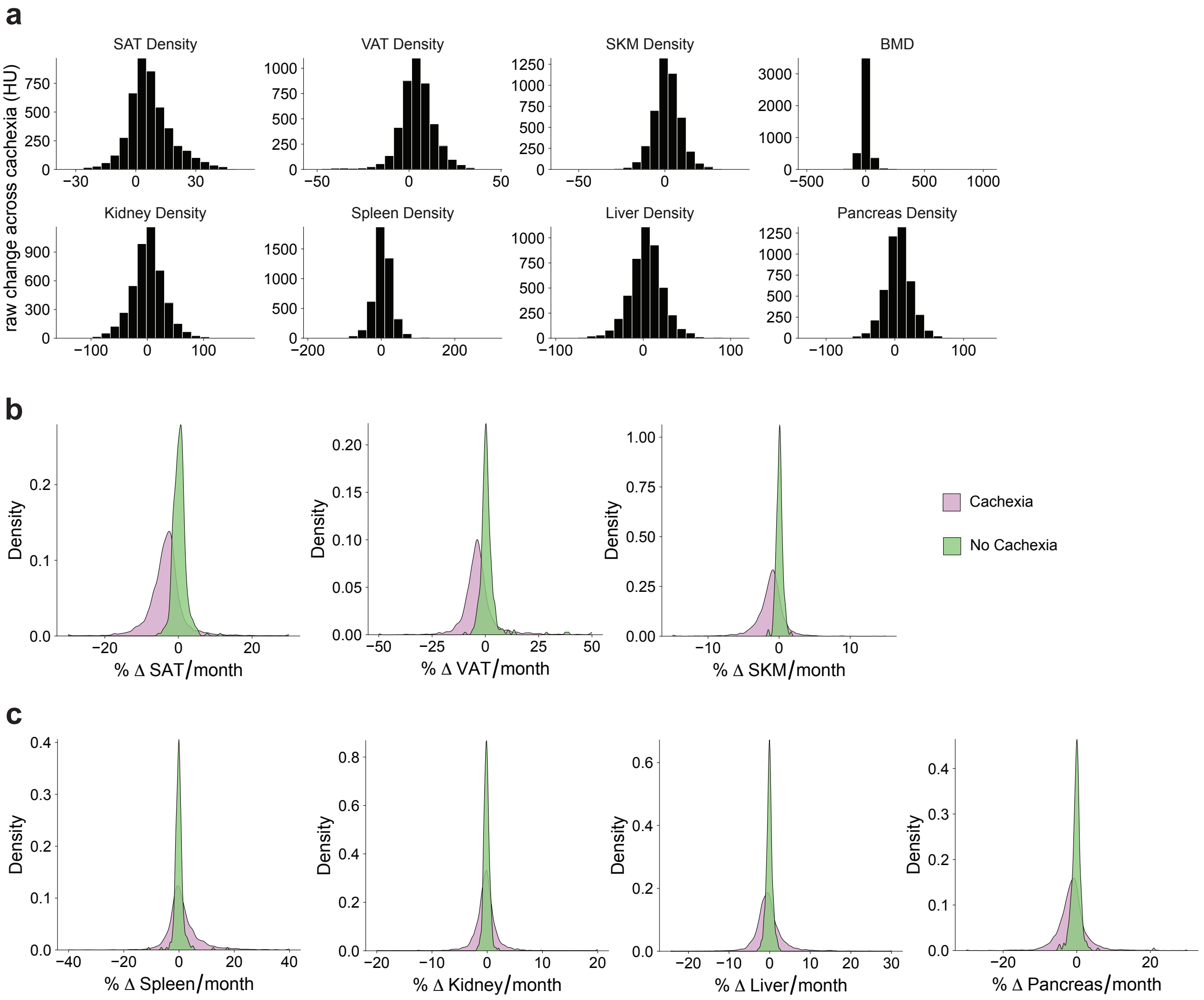


**Supplementary Figure 2: Distribution of body composition changes across cachectic and non-cachectic controls a.** Distribution of densities across body compartments, measured in HU. BMD represents bone mineral density. **b-c.** Density of time-normalized body composition changes in cachectic patients (pink) and in non-cachectic controls (green) across all tissues and organs.

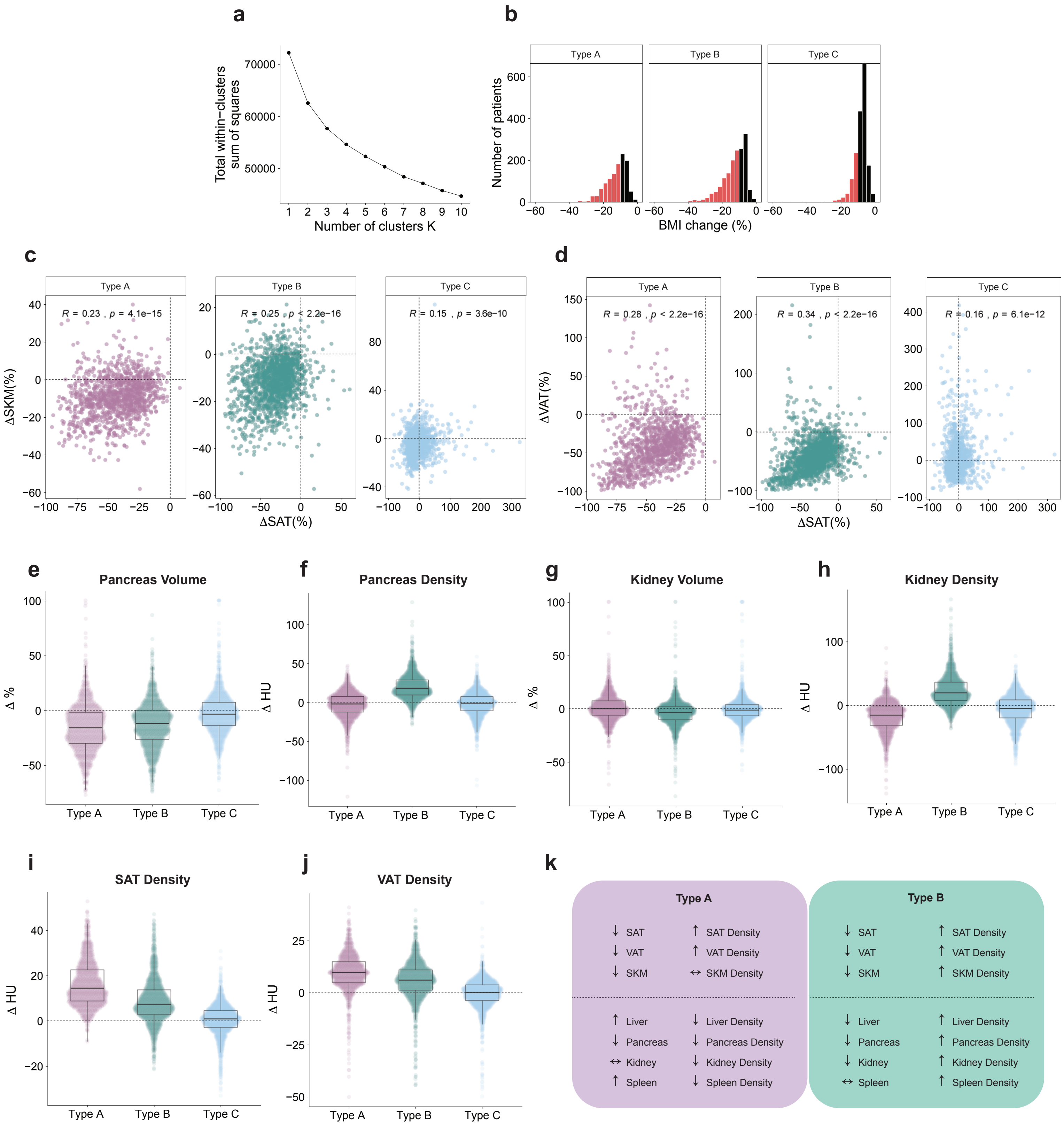


**Supplementary Figure 3: Discovery and characterization of cachexia subtypes. a.** Elbow plot of total within-clusters sum of squares motivating the choice of k = 3. **b.** Histograms of BMI change across the three clusters. Red bars represent changes < -10, while black bars are ≥-10. **c-d.** Correlation of SAT, VAT, and SKM changes across clusters. P and R values derived from Pearson correlations. Dotted lines at 0. **e-j.** Boxplots demonstrating percent and raw changes across organ volumes and densities. Boxplots indicate mean and interquartile range; error bars, s.e.m. The dotted line separates gain and loss at zero. **k.** Summary of body composition changes across Type A/B cachexia.

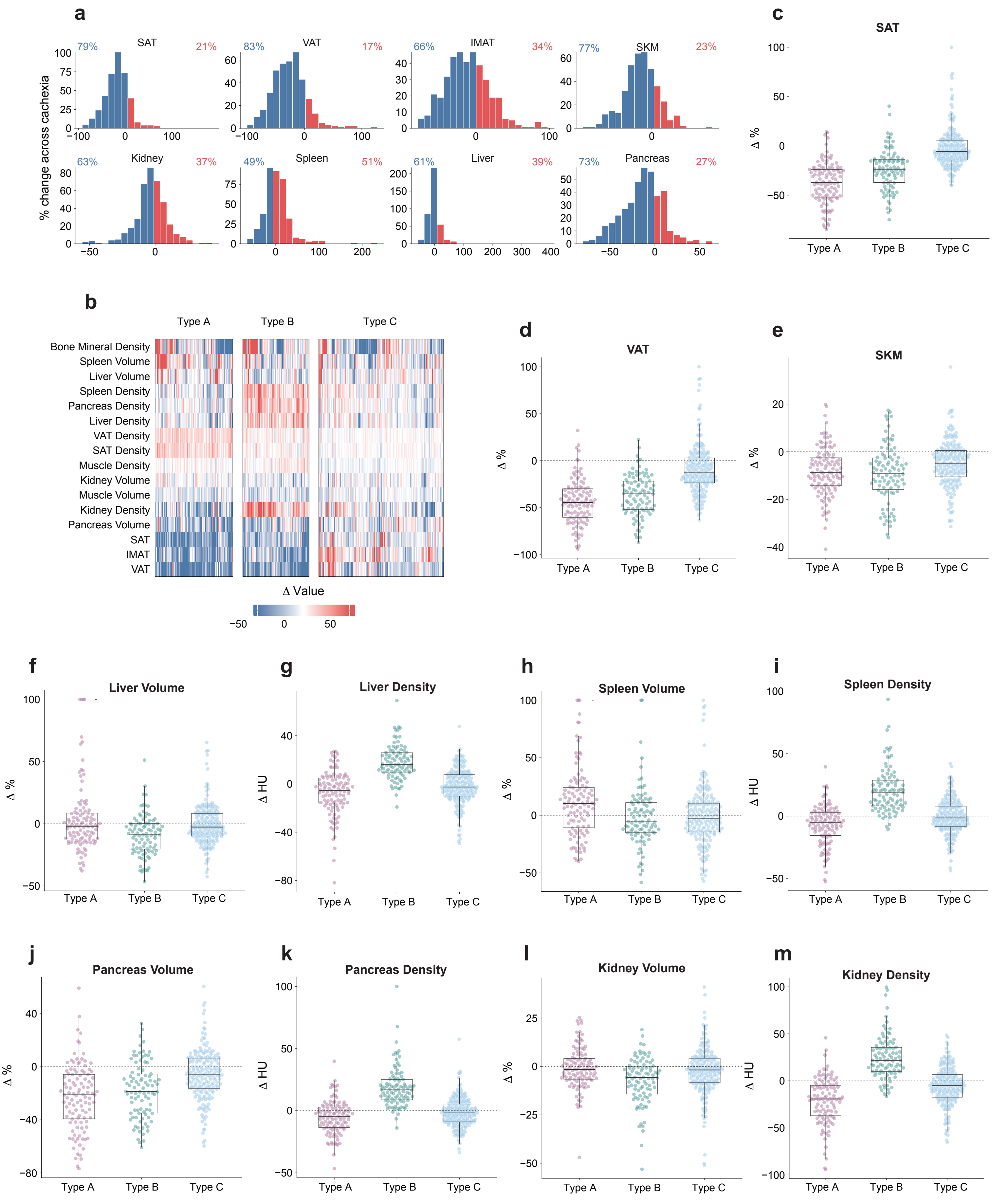
a
% change across cachexia
SAT 79% 21%
VAT 83% 17%
IMAT 66% 34%
SKM 77% 23%
Kidney 63% 37%
Spleen 49% 51%
Liver 61% 39%
Pancreas 73% 27%
b
Type A
Type B
Type C
Bone Mineral Density
Spleen Volume
Liver Volume
Spleen Density
Pancreas Density
Liver Density
VAT Density
SAT Density
Muscle Density
Kidney Volume
Muscle Volume
Kidney Density
Pancreas Volume
SAT
IMAT
VAT
Δ Value
c SAT
d VAT
e SKM
f Liver Volume
g Liver Density
h Spleen Volume
i Spleen Density
j Pancreas Volume
k Pancreas Density
l Kidney Volume
m Kidney Density
Δ %
Δ HU

**Supplementary Figure 4. Subtype discovery in UW validation cohort. a.** Distribution of body composition changes during cachexia across all tissues and organs. **b.** Clustering of body composition changes in UW cohort. **c-m.** Pairwise comparisons of key compartment changes. Boxplots indicate mean and interquartile range; error bars, s.e.m. The dotted line separates gain and loss at zero.

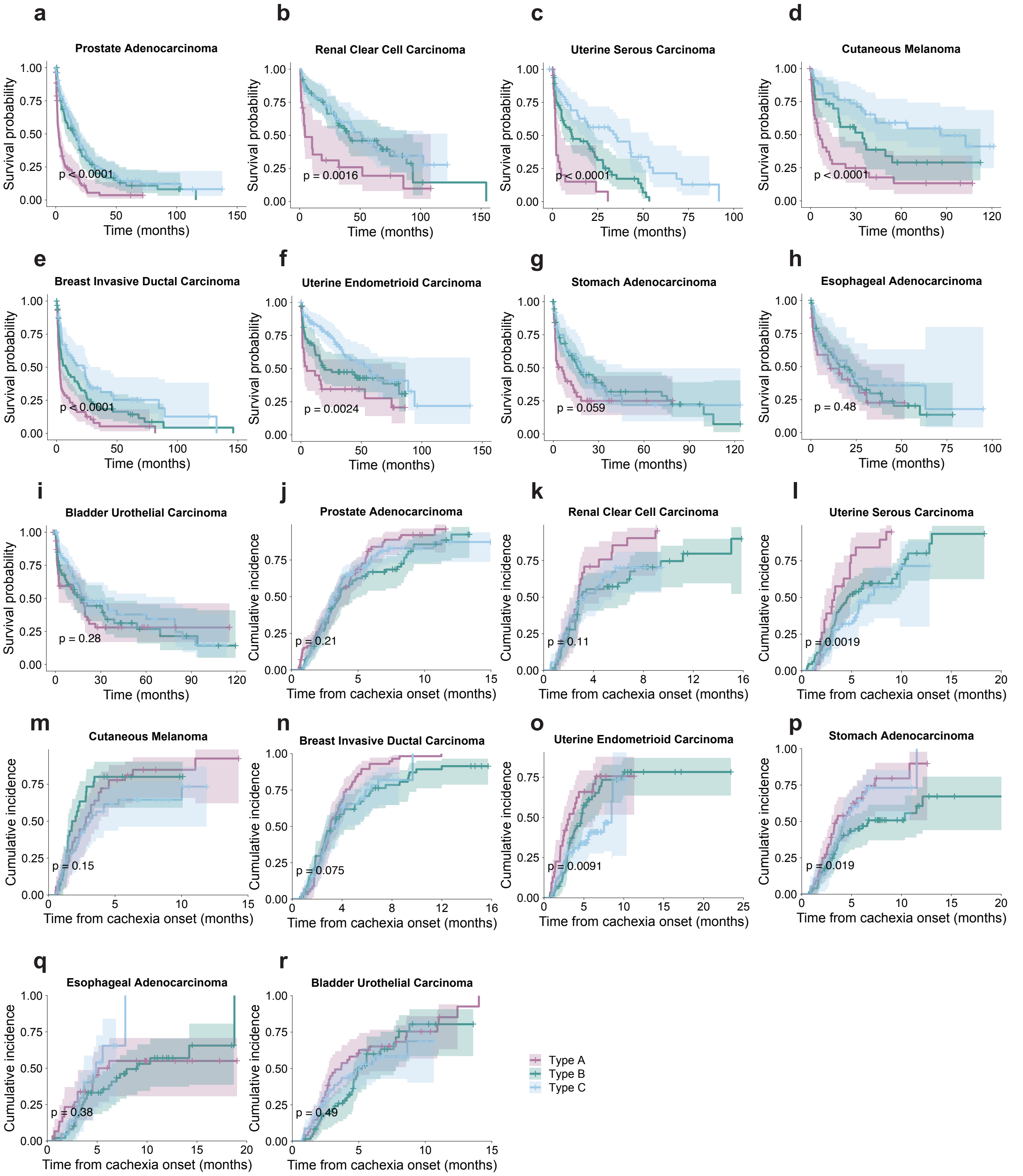


**Supplementary Figure 5: Clinical correlates of cachectic subtypes. a-i.** Kaplan-meier curves modeling survival from cachectic end onwards. **j-r.** Cumulative incidence for first progression event from cachexia onset. P-values from log-rank test.

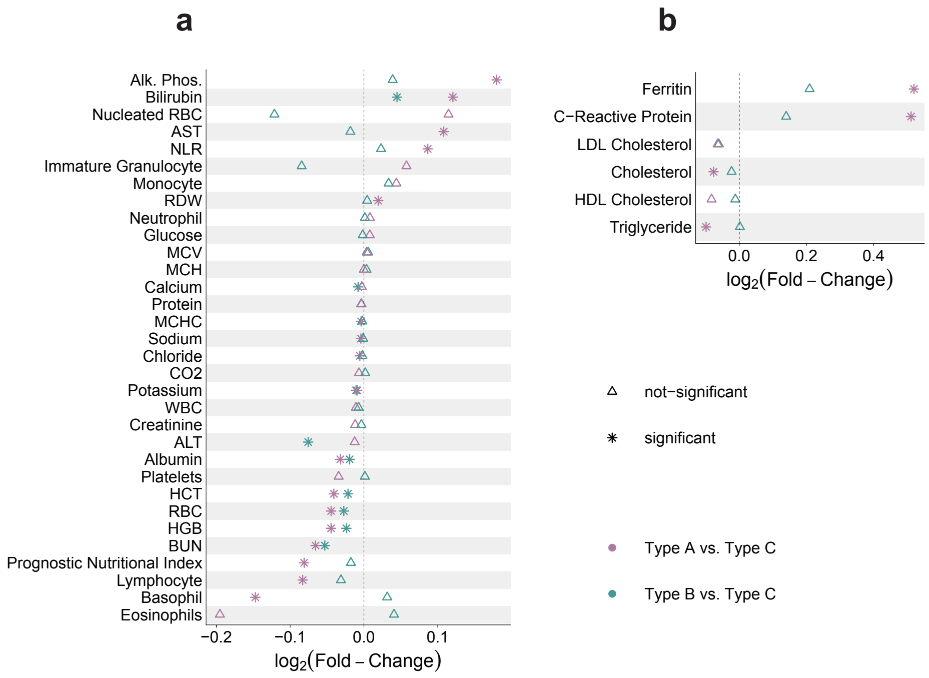


**Supplementary Figure 6: Disease agnostic serological signature of Type A and Type B cachexia. a.** Linear mixed effects models comparing Type A to Type C and Type B to Type C. “*” denotes $q < 0.05$. **b.** Same as a but in the lipid/additionally ordered laboratory panel.
